# Immunohistochemical Profiling of Histone Modification Biomarkers Identifies Subtype-Specific Epigenetic Signatures and Potential Drug Targets in Breast Cancer

**DOI:** 10.1101/2024.12.13.24318980

**Authors:** Zirong Huo, Sitong Zhang, Guodong Su, Yu Cai, Rui Chen, Mengju Jiang, Dongyan Yang, Shengchao Zhang, Yuyan Xiong, Xi Zhang

## Abstract

**Background:** Breast cancer (BC) subtypes exhibit distinct epigenetic landscapes, with triple-negative breast cancer (TNBC) lacking effective targeted therapies. This study investigates histone biomarkers and therapeutic vulnerabilities across BC subtypes. **Methods:** Immunohistochemical profiling of >20 histone biomarkers, including histone modifications, modifiers and oncohistone mutations was conducted on a discovery cohort and a validation cohort of BC tissues, healthy controls and cell line models. Transcriptomic and cell growth analyses were conducted to evaluate the effects of the small-molecule G9a inhibitor in diverse BC models. **Results:** Key histone biomarkers, including H3K9me2, H3K36me2, and H3K79me, were differentially expressed across BC subtypes. H3K9me2 emerged as an independent predictor for distinguishing TNBC from other less aggressive BC subtypes, with elevated expression correlating with higher tumor grade and stage. G9a inhibition impaired cell proliferation and modulated epithelial-mesenchymal transition pathways, with the strongest impact in basal-like TNBC. Disruption of oncogene and tumor suppressor regulation (e.g., TP53, SATB1) was observed in TNBC. **Conclusion:** This study highlights G9a’s context-dependent roles in BC, supporting its potential as a therapeutic target. Findings provide a foundation for subtype-specific epigenetic therapies to improve outcomes in aggressive BC subtypes.

**Clinical Perspective:**

- This study was undertaken to explore the epigenetic landscape of breast cancer subtypes, focusing on histone modifications and their therapeutic potential, particularly through targeting G9a, a histone methyltransferase.
- The study identified subtype-specific histone biomarkers, with H3K9me2 emerging as a key marker distinguishing TNBC from other less aggressive subtypes. G9a inhibition demonstrated robust anti-cancer effects, including cell proliferation impairment and disruption of oncogenic pathways, particularly in TNBC models.
- These findings provide compelling evidence for the development of subtype-specific epigenetic therapies targeting G9a and similar regulators, highlighting their potential to reshape the treatment landscape for TNBC and improve patient outcomes.

## Introduction

Histones, as the core components of chromatin, undergo chemical modifications that regulate gene expression and are intimately linked to cancer development[1]. Histone modifications such as methylation and acetylation act as pivotal regulators of gene activity, influencing transcriptional regulation and tumor progression depending on their type and genomic context[2, 3]. For instance, H3K4me2 is a mark of active transcription, typically found at promoters of genes primed for expression. Conversely, H3K9me2 is associated with transcriptional repression and heterochromatin formation, silencing genes that may drive tumorigenesis. However, the loss of H3K9me2 can paradoxically activate oncogenes, contributing to cancer progression.

BC is a highly heterogeneous disease, classified into distinct molecular subtypes based on the expression of classic biomarkers such as Estrogen Receptor (ER), Progesterone Receptors (PR) and Human Epidermal Growth Factor Receptor 2 (HER2)[4]. Among these, TNBC poses the greatest clinical challenge due to its aggressive nature and the absence of well-defined therapeutic targets. Bridging the treatment gaps in TNBC necessitates innovative approaches, particularly those focusing on its unique molecular and epigenetic characteristics[5]. Aberrations in histone modifications in cancer can vary significantly, even within the same cancer type. Research has demonstrated that different molecular subtypes of BC (e.g., luminal A and TNBC) exhibit distinct patterns of histone modifications, reflecting subtype-specific epigenetic landscapes[6]. Although evidence from cell line studies have highlighted that TNBC is the molecular subtype that most closely associated with distinct histone modification patterns[7, 8], research focusing on human tissue remains limited. This gap underscores the need for studies using patient-derived samples to validate findings from in vitro models and to explore the clinical relevance of these epigenetic alterations in tumor progression.

The unique histone modification patterns of TNBC, once fully characterized, may distinguish it from subtypes like luminal A and HER2-enriched breast cancers, highlighting new possibilities for subtype-specific therapies[9]. Current epigenetic treatments, including DNA methyltransferase (DNMT) and histone deacetylase (HDAC) inhibitors, show clinical promise[10]. Histone methyltransferase (HMT) inhibitors correspond to the third generation of epigenetic drugs capable of writing or deleting epigenetic information[11, 12]. EZH2 inhibitor, targeting H3K27me3, is the first and only HMT inhibitor that approved by FDA in 2020 for the treatment of certain cancers[13, 14]. Emerging targets like H3K79me1/2/3 (DOT1L inhibitors) and H3K9me2 (G9a inhibitors) are showing potential in preclinical studies of breast cancer[15–17].

In this study, the global expression of histone modifications, histone modifiers, and oncohistone mutations was characterized in BC tissues using immunohistochemical (IHC) staining. Expression patterns were validated across two independent cohorts and compared to those in healthy tissue samples and BC cell lines. Subtype-specific histone modifications were identified, with several significant ones showing strong associations with the TNBC subtype, tumor grade, and stage. Treatment with a small-molecule epigenetic inhibitor significantly impaired cell growth in both estrogen receptor-positive and -negative cell lines. Transcriptomic and pathway analyses further confirmed distinct responses between the two cell line types. Together, these findings highlight specific histone modifications as critical markers and drivers of advanced or accelerated breast cancer progression and establish histone modifiers as promising therapeutic targets in BC.

## Method

### Study Design

To identify and evaluate the potential of 21 pathological biomarkers in breast cancer, this study used tissue microarray technology, allowing for high-throughput molecular analysis of 196 tissue samples from a discovery cohort of 98 breast cancer patients (Fig. 1). The IHC profiling results of TNBC were then validated by additional 20 tissue blocks from a validation cohort of 20 TNBC patients. Eligible participants of both cohorts were women aged 35-70 years with a first primary diagnosis of breast cancer (Table 1). The distributions of clinical characteristics and traditional pathological biomarker results are shown for both the discovery and validation cohorts, as provided by the commercial vendors of BC tissues. Based on IHC assessments of ER, PR, HER2, and Ki-67, breast cancer samples in this study were classified into 79 luminal A cases, 53 luminal B cases, 23 HER2-enriched cases, and 41 TNBC cases in the discovery cohort, as well as 20 TNBC cases in the validation cohort. Histone biomarker signatures identified from tissue samples were subsequently evaluated in BC cell lines. To explore their functional relevance, the selected HMT inhibitor, G9a inhibitor UNC0642, was applied to assess its effects on cell proliferation, impacted gene expression and signaling pathways. All study procedures were approved by the Institutional Ethics Committee and the Institutional Review Board of Northwest University (approval number: 200,402,001).

**Figure 1.**
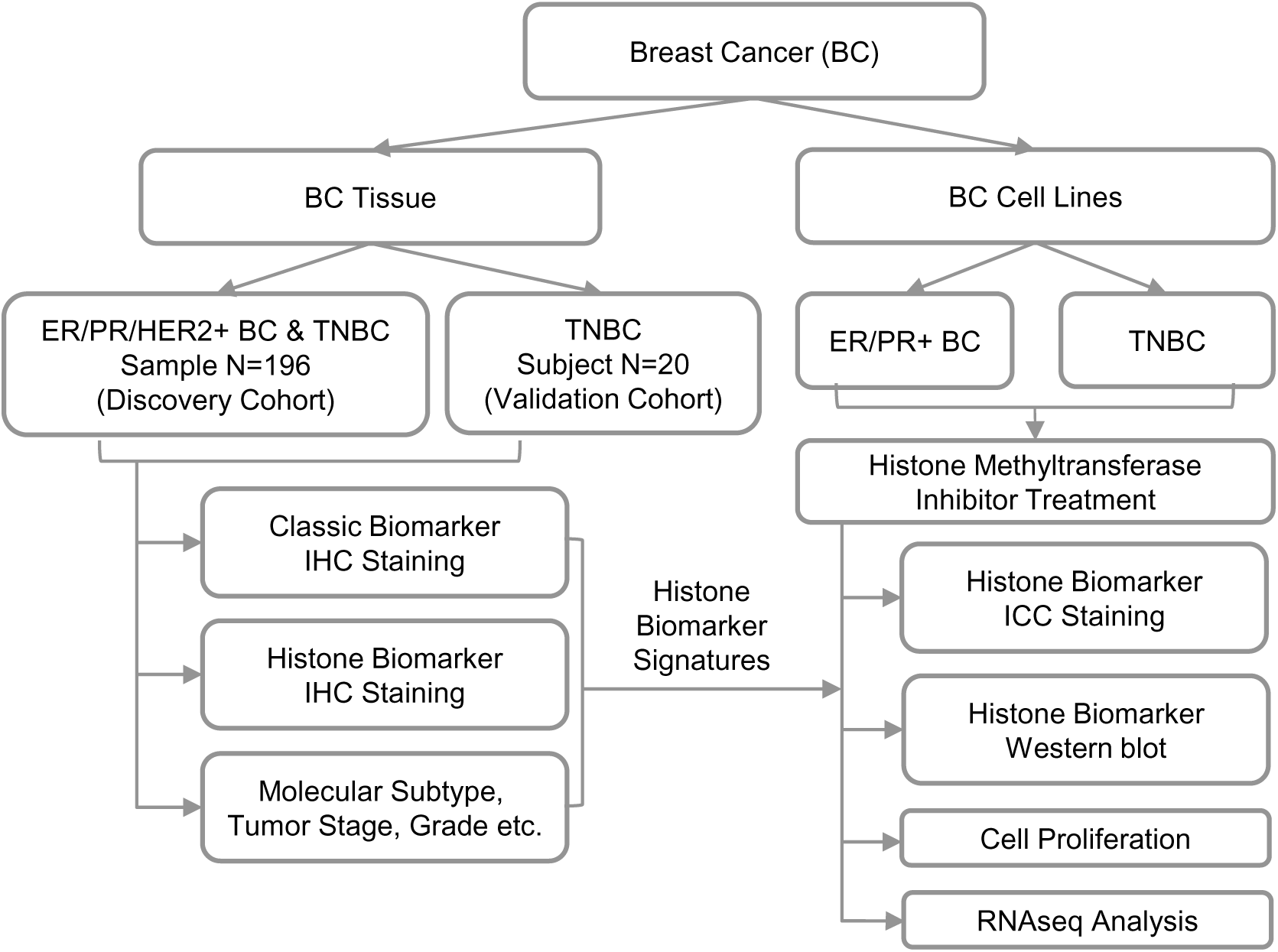

### Tissue Specimens

We collected 216 tissue samples from commercial sources for this study. A high-density breast cancer tissue microarray (chip number F1961101 B06-F1961101 B25) was purchased from Xi’an Bioaitech Co., Ltd. Each chip included two 1-mm tissue cores from 98 patients with various molecular subtypes, forming a discovery cohort of 196 samples. A validation cohort was established using 20 TNBC tissue slides procured from Shanghai Xinchao Co., Ltd. (sample codes listed in Table S4). Clinical characteristics and biomarker data were available for all tissue samples (Table 1).

### Cell Culture

Three breast cancer cell lines—MCF-7 (luminal A), MDA-MB-231 (claudin-low TNBC), and MDA-MB-468 (basal-like TNBC)—and two T-lymphocytic leukemia cell lines (HPB-ALL and LOUCY) were used in this study. Origin and catalogue number of cell lines are listed in Table S4. Three BC cell lines were cultured in Dulbecco’s modified Eagle’s medium (DMEM). HPB-ALL and LOUCY cells were cultured in RPMI-1640. In both media, 10% fetal bovine serum and 1% penicillin/streptomycin were added. The above cells were grown in a humidified 5% CO_2_ incubator at 37°C. For estrogen treatment, cells were starved in phenol red-free DMEM supplemented with 10% charcoal-stripped FBS for 2 days, and 73.4 mM estrogen were added to the medium (3 and 6 h for gene expression studies, and 6 h for immunocytochemical (ICC) experiments) before the cells were collected. The detailed list of reagents and kits used in the method can be found in Table S4.

### IHC and ICC

IHC analysis was performed using the streptavidin-biotin-peroxidase complex method to evaluate 20 histone biomarks (including H3) of interest. Tissue chips and slides were processed by deparaffinization, antigen retrieval, blocking with goat serum, and incubation with primary antibodies overnight at 4°C. Secondary antibodies and DAB were used for detection, followed by counterstaining and mounting. Anti-H3 served as a control. Details of antibodies, reagents and kits are provided in Table S4. ICC was performed using the same principles as IHC. Cells were fixed in 4% paraformaldehyde, permeabilized with 0.25% Triton X-100, and blocked with 10% goat serum. Primary antibodies were incubated overnight, followed by secondary antibody incubation and DAB detection.

### Cell Viability and Proliferation Assay

Cell viability was assessed using the Cell Counting Kit-8 (CCK-8). Cells were seeded in 96-well plates (5 × 10³/well), treated with UNC0642 or DMSO (Control), and incubated. After treatment, 10 µL CCK-8 solution was added, and OD450 was measured. For proliferation assays, cells were treated with 2 µM UNC0642 or DMSO and analyzed over six days using the same method. The detailed list of reagents and kits used in the method can be found in Table S4.

### Western Blotting

Proteins form BC cells, which were treated with 2 µM UNC0642 or DMSO for 48 h and were extracted using RIPA buffer, quantified using the BCA assay, and separated by SDS-PAGE. After transfer to PVDF membranes, blots were incubated with primary and secondary antibodies. Signals were detected using enhanced chemiluminescence, with H3 as a loading control. Antibodies, reagents and kits details are listed in Table S4.

### RNA isolation and Sequencing

Total RNA from BC cells collected after 4 days of treatment with 2 µM UNC0642 or DMSO was extracted using Trizol and assessed for purity and integrity. RNAseq libraries were prepared using the TruSeq RNA Sample Prep Kit and sequenced on an Illumina NovaSeq 6000 platform. Clean reads were analyzed using STAR aligner and DESeq2 to identify differentially expressed genes (DEGs). Genes in breast cancer cell lines that displayed at least one-fold difference in gene expression between comparison groups (fold change > 1 or < -1, FDR p < 0.05) were considered significant DEGs and carried forward in the analysis. Pathway and gene ontology (GO) enrichment analysis was performed via an integrated platform KOBAS 3.0 and GSEA using MSigDB gene sets within selected collections H, C2, C4, C6. Bioinformatics analysis was performed and visualized using R version 3.6.1 or Python version 3.7.9. The detailed list of reagents and kits used in the method can be found in Table S4.

### Real-time quantitative PCR (RT-qPCR)

Real-time quantitative PCR was performed using Bio-Rad real-time PCR systems. Total RNA (1 µg) was reverse transcribed into cDNA using the Hifair III 1st Strand cDNA Synthesis SuperMix for qPCR. mRNA expression levels were quantified using Hieff® qPCR SYBR Green Master Mix, with relative expression calculated by the 2^−ΔΔCt^ method. Each sample was tested in triplicate, and primers, reagents and kits are listed in Table S4.

### IHC Evaluation and Cutoff Determination

Histone expression was quantified using integrated optical density (IOD) values from Image J (Media Cybernetics Inc., Rockville, MD, USA) analysis[18]. Receiver operating characteristic (ROC) curve analysis was used to determine cutoff scores for each histone, distinguishing high and low expression[19]. Clinical features, including age, grade, T stage, N stage, tumor stage, and receptor status, were classified for analysis. For each histone biomarker, the point of maximum sensitivity and specificity was selected based on the area under the curve. Samples were classified as low expression if their integrated optical density was below the cutoff and high expression if equal to or above the cutoff. Clinical features used for stratification in ROC analysis included: Age (≥ 50 vs. <50 years), Tumor Grade (Grade 2 vs. 2-3/3), T stage (T1/T2 vs. T3/T4), N stage (N0 vs. N1/N2), Tumor Stage (Stage I vs. Stage II/III), Receptor Status (ER/PR/HER2 negative vs. positive), Ki-67 Index (<14% vs. >14%). These cutoff values provided a basis for categorizing histone expression and analyzing its association with breast cancer subtypes and clinical outcomes.

### Statistical Analysis

Statistical analyses were conducted using SPSS (v13.0) and GraphPad Prism 9. T-tests, chi-square tests, and logistic regression were used to analyze data.

## Results

### Pathological Scoring Identified Key Histone Biomarkers across BC Molecular Subtypes

To characterize the global expression of histone biomarkers in tissue samples, IHC staining was initially performed on two cohorts of BC patients. The analysis included 12 histone modifications, 7 histone modifiers, 2 oncohistone mutations, and histone H3 as a control signal (Fig. 2A). The histone modifications and modifiers selected in this study are recognized for their roles in driving dysregulated gene expression patterns in breast cancer (Fig. 2A) and will henceforth be termed histone biomarkers. The first cohort, referred to as the discovery cohort, comprised 196 tissue spots (Fig. 2B). These were derived from 98 patients (Table 1), with duplicate spots collected per patient. Each IHC staining spot was scored by combining the area and density of the dyed region assessed by Image J. The samples were then categorized into two classes: High-IHC score and Low-IHC score (Fig. 2C).

**Figure 2.**
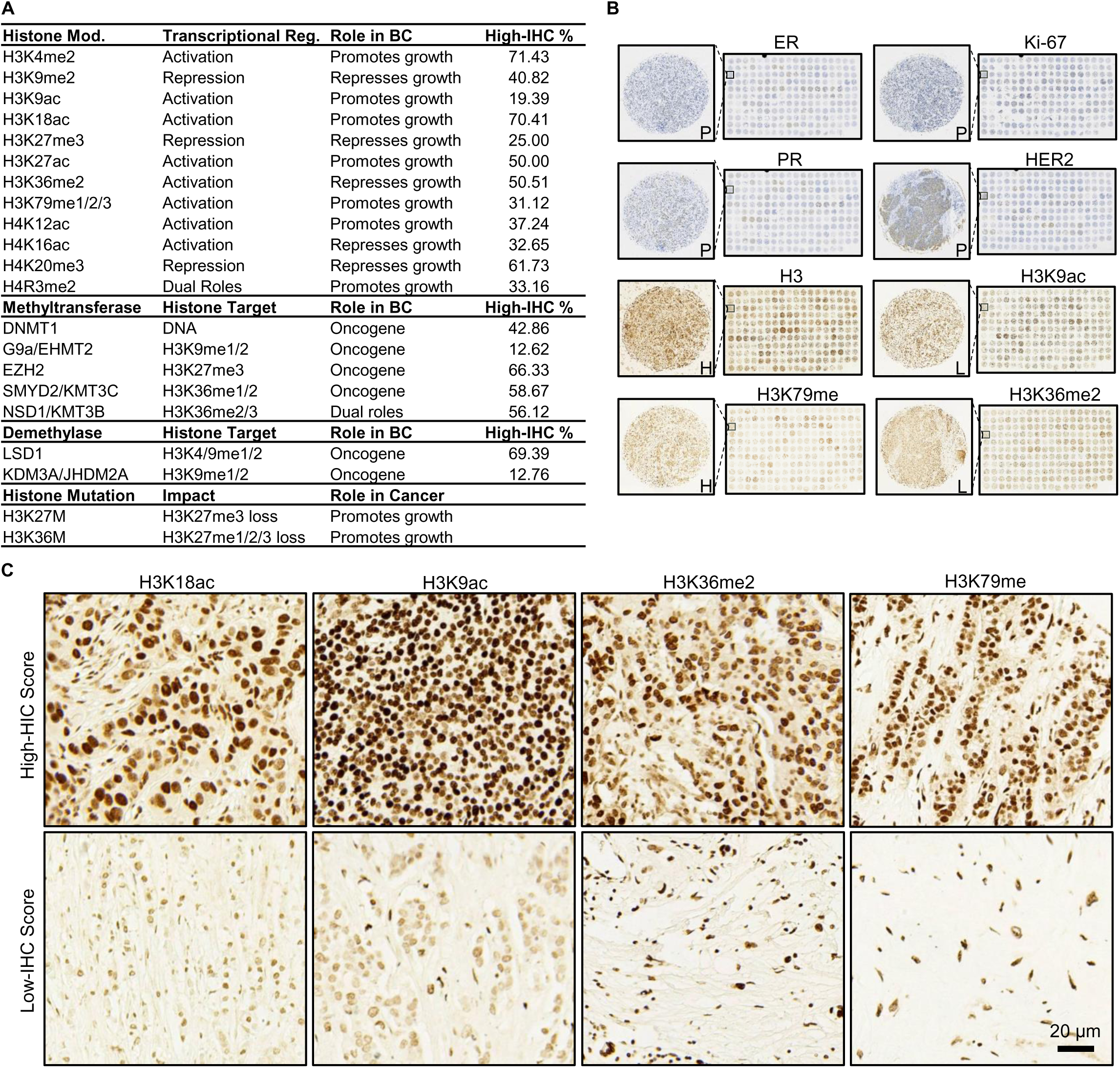

The molecular subtypes of BC tissues (luminal A, luminal B, HER2-enriched, and TNBC) in the discovery cohort were determined by analyzing IHC staining patterns for ER, PR, HER2, and Ki-67 (Fig. 3A). Heatmap of high-IHC % suggests that high-IHC scores are not uniformly distributed among the molecular subgroups (Figs. 3B & 3C). Statistical analyses were conducted to assess the significance of the distribution of histone biomarker levels. Initially, chi-square tests were performed to assess the difference in the distribution of histone biomarkers across sample subgroups defined by clinical characteristics as described in Table 1. As presented in Table 2 and Fig 3B, six histone biomarkers (H3K4me2, H3K9me2, H3K36me2, LSD1, NSD1, and DNMT1) showed significant differences across the four molecular subtypes. H3K9me2, H3K36me2, H3K79me (me1/2/3) and two acetylation markers (H3K18ac and H4K16ac) exhibited significant differences in TNBC compared to other molecular subtypes.

**Figure 3.**
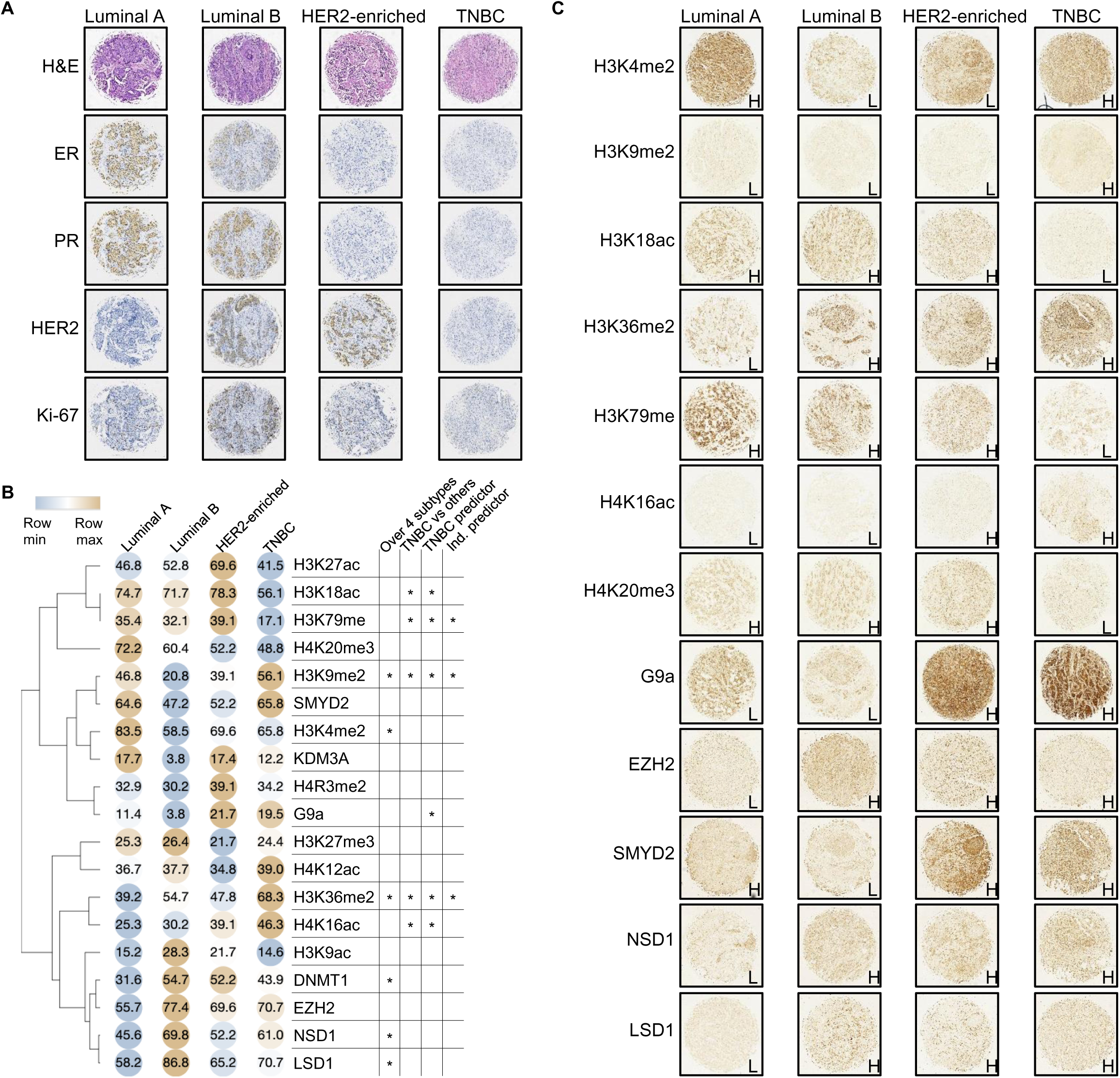

Logistic regression analysis was then conducted to explore the relationship between IHC scores (High-IHC vs. Low-IHC) and histone biomarkers (Table 3), excluding the oncohistone mutations that were not detectable in any of the tissue samples (Fig. S1). Univariate regression was performed to evaluate the association of each histone biomarker with the IHC score individually. Histone biomarkers with p-values below 0.05 in the univariate analysis were subsequently included in the multivariate regression model. The analysis highlighted H3K9me2, H3K36me2, and H3K79me (me1/2/3) as independent predictors of altered IHC scores in TNBC compared to other molecular subtypes. G9a, the only histone modifier that exhibited significant differences in TNBC, was determined to be a dependent predictor. Furthermore, H3K9me2 and H3K36me2, along with H3K4me2 and others, were also identified as independent predictors for the luminal A and luminal B subtypes.

### Key Histone Biomarkers Serve as Indicators of Advanced Tumorigenesis in BC

Next, we aimed to evaluate the expression levels of selected histone biomarkers in adjacent healthy breast tissue compared to breast cancer tissue. Due to material limitations, the analysis focused on histone H3, three specific histone modifications (H3K4me2, H3K9me2, and H3K9ac), and two corresponding modifiers (G9a and LSD1). Representative IHC images of luminal A breast cancer and TNBC are displayed side by side (Fig. 4A). IHC images of two additional independent TNBC predictors, H3K36me2 and H3K79me, are presented alongside LSD1. Histone biomarkers and corresponding modifiers exhibited clustered expression patterns, as demonstrated in heatmap analyses (Fig. 3B). Interestingly, histone biomarkers commonly associated with transcriptional repression (H3K9me2, G9a and LSD1) were more highly expressed in TNBC compared to luminal A and were also elevated in breast cancer tissue compared to healthy tissue. In contrast, the expression of biomarkers associated with transcriptional activation (H3K4me2 and H3K9ac) followed a decreasing trend, with the lowest levels observed in TNBC, intermediate levels in luminal A, and the highest levels in healthy tissue.

**Figure 4.**
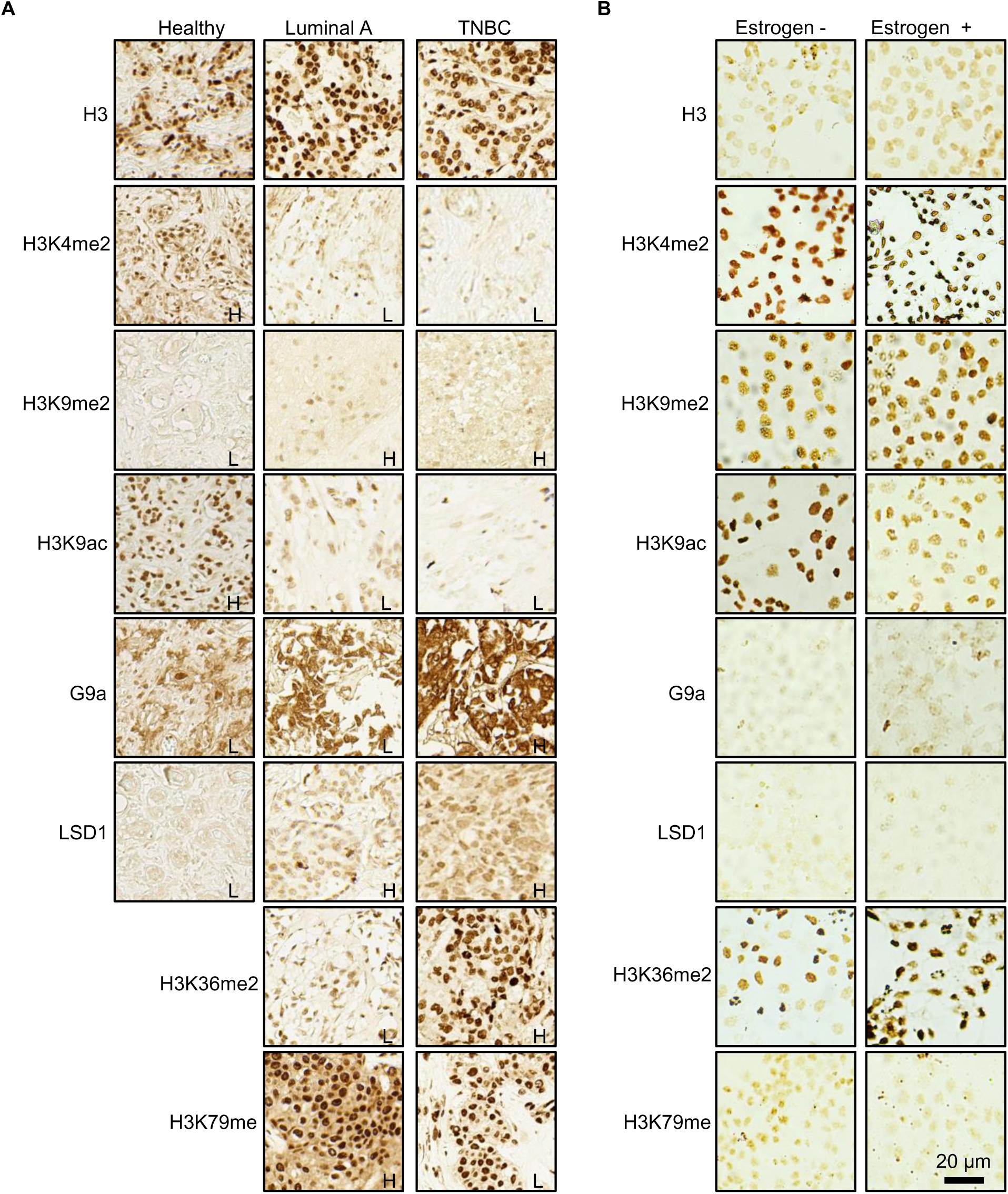

The MCF-7 cell line treated with estrogen is a well-established luminal A breast cancer model for studying estrogen-driven regulation. A positive transcriptional response to estrogen is central to the biology of ER-positive breast cancer and serves as a hallmark of disease progression and treatment response (Fig. S2). To further explore the identified key histone biomarkers, their global expression was assessed via ICC staining in estrogen-treated MCF-7 cells. Notably, the histone biomarkers H3K9me2 and G9a, out of the five markers characterized above in tissue samples, showed increased expression in estrogen-treated MCF-7 cells compared to estrogen-deprived cells (Fig. 4B). Additionally, several other methylation and acetylation markers exhibited altered expression in this cellular model (Fig. S3), indicating a complex epigenetic landscape in ER-responsive regulation in breast cancer. These findings reinforce the association of H3K9me2 and G9a with advanced tumorigenesis in breast cancer and underscore their roles in promoting estrogen-dependent tumor growth and progression.

### Validation of Altered Global Histone Modification in TNBC Tissue

As the role of G9a and H3K9me2 in ER-positive breast cancer is better understood, the focus of this study shifted toward further exploring the significance of key histone biomarkers in TNBC. To validate these findings, a second breast cancer cohort was established, comprising 20 tissue slides from 20 TNBC cases. The clinical characteristics of the validation cohort are summarized, with no significant differences observed between the discovery and validation cohorts (Table 1). Each tissue sample was stained for 4 histone biomarkers, and the IHC results were scored and classified into IHC-high and IHC-low categories as described previously (Table 4). The IHC-high percentages of TNBC-independent predictors (H3K9me2, H3K36me2, H3K79me) and modifier G9a in the validation cohort are provided in Table 4, alongside corresponding metrics for TNBC and luminal A cases in the discovery cohort. Significant differences (p<0.05) were observed in the IHC-high% of all three independent predictors when comparing the at least one TNBC cohort to luminal A, but no significant difference was found between validation TNBC and discovery TNBC. This finding confirmed the agreement on the significance of key histone biomarkers between two TNBC cohorts and further highlighted the distinction between TNBC and luminal A, in addition to the results described above comparing TNBC with all non-TNBC subtypes.

Among the three histone biomarkers validated in this study, H3K9me2 was selected for further analysis. One reason for this selection is that while the roles of G9a and H3K9me2 have been extensively studied in ER-positive breast cancer, their involvement in TNBC remains largely unexplored, despite strong prior associations. Another reason is that global H3K9me2 signals were significantly linked to tumor grade, as well as T and N stages (Table 2, Fig 5A). Notably, a gradual increase in H3K9me2 level (IHC-high%) was observed with higher tumor grade or stage (T4 > T3 > T2) (Fig 5B). This pattern suggests that H3K9me2 may be associated with more aggressive or advanced breast cancer status or subtypes, such as TNBC, which are characterized by higher proliferation rates and poorer clinical outcomes.

**Figure 5.**
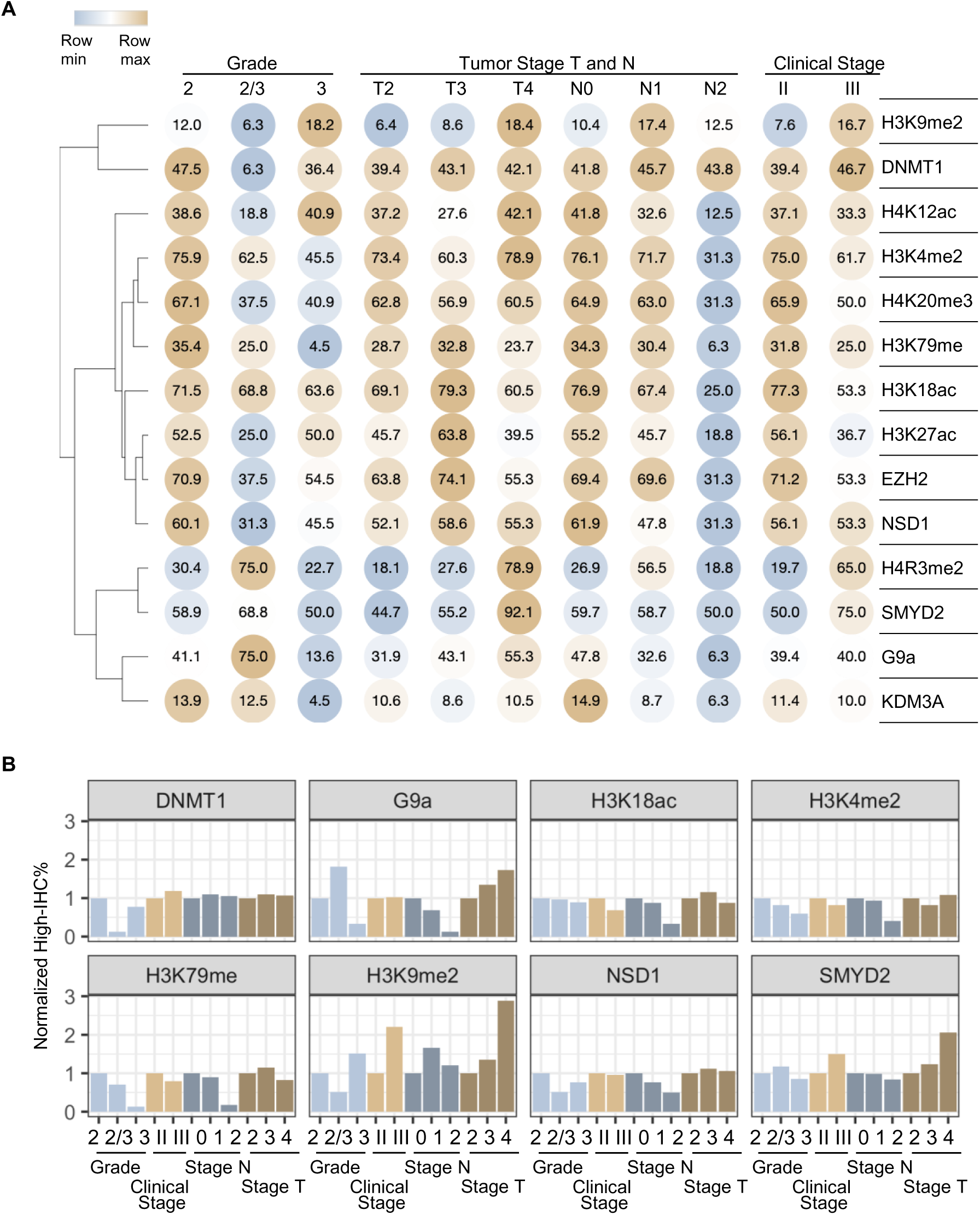

### G9a Inhibition Disrupts Proliferation and Signaling Pathways in Breast Cancer Cell Lines

UNC0642, a novel small-molecule inhibitor targeting the catalytic activity of G9a, was evaluated in MCF-7 (luminal A), MDA-MB-231 (claudin-low TNBC), and MDA-MB-468 (basal-like TNBC) cell lines. Cell viability assays were performed to optimize the dose and treatment duration (Fig. 6A). Finally, non-cytotoxic concentrations of UNC0642 (2 µM) were applied in all subsequent experiments. This inhibitor demonstrated significant efficacy in reducing H3K9me2 levels in all three cell lines, as confirmed by both pathological staining and Western blotting analysis (Figs. 6B & 6C). Other histone modifications, such as H3K4me2, or alternative modifications at the same residue, such as H3K9ac, remained unaffected, indicating the high specificity of UNC0642. Interestingly, G9a protein levels were also reduced in basal-like TNBC cells, which exhibited a more pronounced decrease in H3K9me2 compared to claudin-low TNBC cells. UNC0642 treatment resulted in marked growth inhibition across all tested cell lines, with the most significant effects observed in basal-like TNBC cells, followed by claudin-low TNBC cells, and moderate effects in luminal A cells (Fig. 6D).

**Figure 6.**
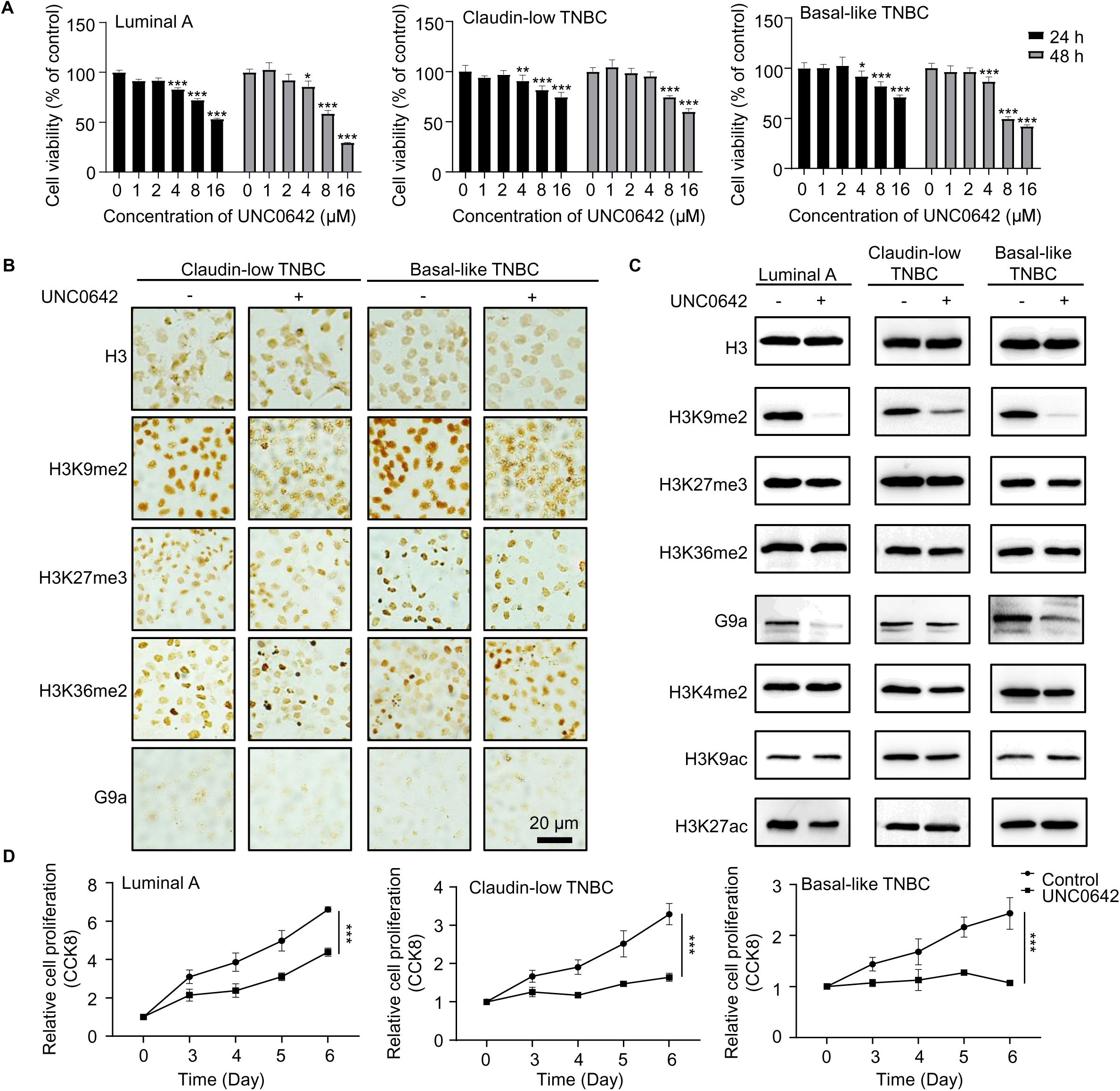

In line with the expected effects of losing the repressive histone mark H3K9me2, the number of up-regulated DEGs was 5-fold higher than down-regulated DEGs in two ER-negative cell lines (Fig. 7A, Fig S4A, Table S1). In contrast, ER-positive MCF-7 cells showed a balanced number of up- and down-regulated DEGs, aligning with prior reports that G9a mediate a direct methylation on ER that was functionally linked to breast cancer progression [17, 20] (Fig. 7A, Fig S4A, Table S1). To further understand the impact of UNC0642, we performed pathway, GO, and GSEA analyses to evaluate gene group and pathway-level responses. The results revealed both shared and distinct transcriptional responses across the three cell lines, highlighting subtype-specific mechanisms of action for the inhibitor (Fig. 7B, Fig S4A). In MCF-7 cells, estrogen-responsive pathways and gene sets were exclusively down-regulated, supporting previous findings that G9a acts as an ERα coactivator [17] (Fig. 7C, Fig. S4B, Tables S2 & S3). Interestingly, G9a inhibition in MDA-MB-231 cells led to a transcriptional response characterized by up-regulation of oncogenes such as SREBF and down-regulation of tumor suppressor genes such as TP53, STAT3 and SATB1 (Fig. 7D, Tables S2 & S3). Cell proliferation pathways and extracellular matrix (ECM)-associated gene sets are consistently down-regulated in both ER-negative and ER-positive cells, emphasizing the shared growth-suppressive effects of G9a inhibition and underscoring its significant therapeutic potential across these breast cancer subtypes (Fig. 7E).

**Figure 7.**
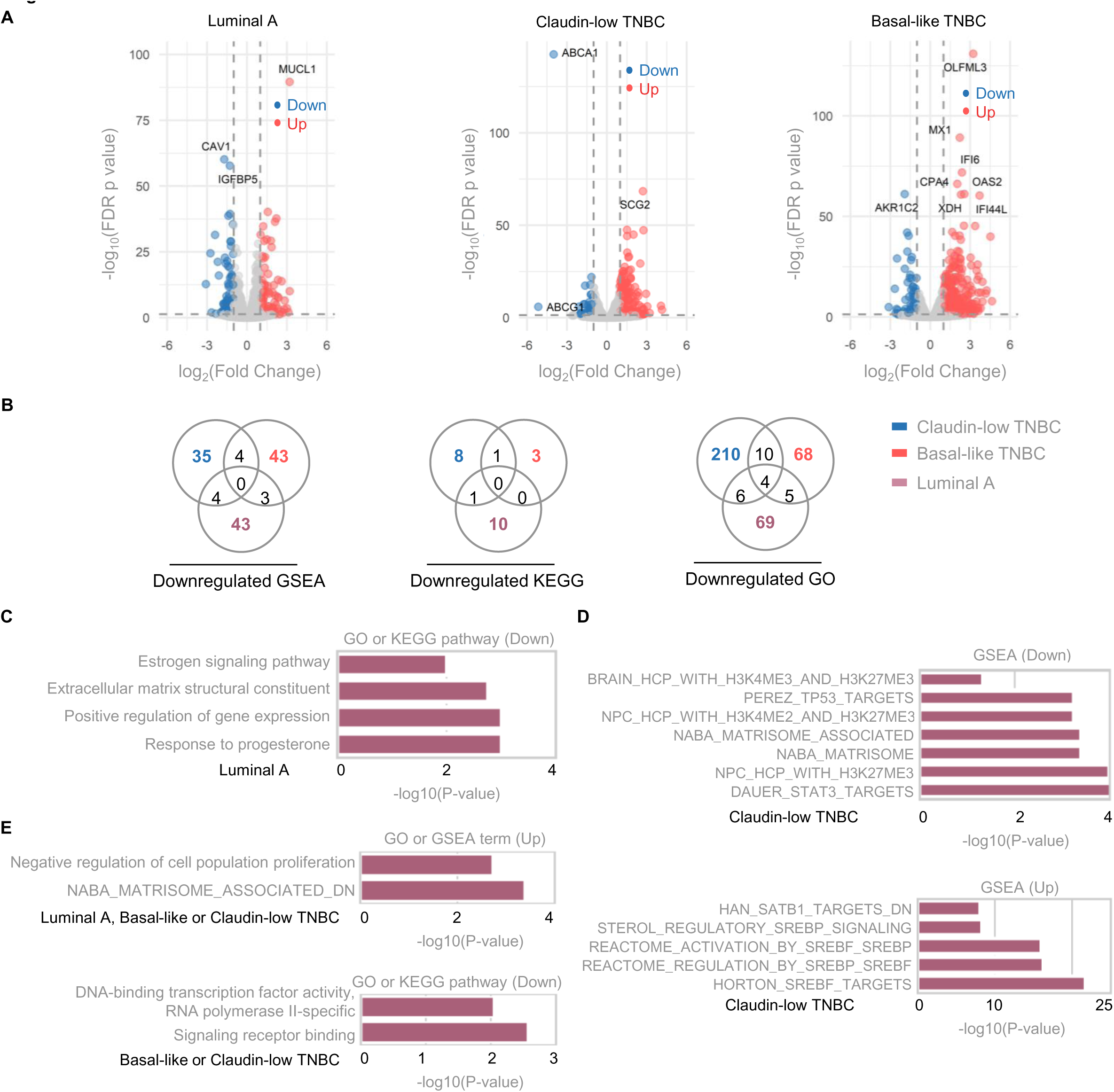

## Discussion

Significant efforts have been dedicated to systematically profiling histone modifications and the enzymes that regulate them in ER-positive BC[21]. These studies have been pivotal in driving the development and clinical application of the first two generations of epigenetic drugs, such as DNMT inhibitors and HDAC inhibitors, with the third generation targeting HMTs now emerging. However, similar insights into histone modification landscapes and their therapeutic potential remain scarce for TNBC, a subtype known for its aggressive and invasive nature. A greater challenge in developing epigenetic drugs is their specificity, which requires emphasis on identifying biomarkers capable of predicting how individual patients will respond to these therapies. This study aimed to address this critical knowledge gap by unveiling key histone biomarker patterns in BC with a specific focus on TNBC. Among the >20 histone biomarkers evaluated, our findings reveal significant alterations in 4 specific histone modification/modifier pairs (H3K4me2/LSD1, H3K9me2/G9a, H3K36me2/NSD1, DNMT1) across the four major BC molecular subtypes. These findings align with the established evidence supporting epigenetic therapy in driving ER-positive breast cancer progression[11, 13]. For example, LSD1 was proposed as a therapeutic target, not only as a standalone approach but also in combination with hormonal therapies[22]. Given the availability of LSD1 inhibitors in preclinical and clinical stages, our results further support their exploration as a promising additional therapy in breast cancer[14, 23].

In our analyses with TNBC cohorts, three histone methylation signals (H3K9me2, H3K36me2, and H3K79me) emerge (and were validated) as independently associated with TNBC, underscoring their potential role as subtype-specific epigenetic markers and therapeutic targets. This result aligns with published reports emphasizing the roles of methylations on H3K36 and H3K79 in TNBC epigenetic regulation. For instance, distinct patterns of H3K36me3 have been observed in TNBC cell lines, particularly in relation to androgen receptor pathway activity[7]. Additionally, the chromatin state marked by H3K4me3 and H3K79me2 at loci such as AFAP1-AS1 has been identified as a hallmark of TNBC, particularly in driving active transcription programs in these cells[7]. Independent studies have confirmed that both NSD3, a histone methyltransferase responsible for H3K36me2, and DOT1L, which methylates H3K79, are highly expressed in the MDA-MB-231 TNBC cell line[24, 25]. Inhibition or knockdown of these histone writers leads to a reduction in H3K36me2 and H3K79me levels, respectively. This reduction in histone modifications is associated with a significant decrease in cell proliferation, migration, and invasion. Notably, in the context of TNBC, these effects are often linked to the reversal of epithelial-to-mesenchymal transition (EMT), a key process that contributes to increased metastatic potential and the aggressive nature of cancer cells[26]. Here, our findings provide direct evidence from multiple cohorts of human tissue to support observations previously made in cell line studies and underscore the therapeutic potential of targeting histone writers like NSD3 and DOT1L in TNBC treatment strategies.

Mutations in histone H3 (such as H3K36M and H3K27M), so-called “oncohistones”, have been identified as significant contributors to tumorigenesis in certain cancers and thus was investigated in this study[27]. H3K36M is known to drive skeletal tumors like chondroblastoma by dominantly inhibiting H3K36 methylation, leading to transcriptional dysregulation and altered differentiation[28]. The H3K36M oncohistone primarily inhibits several H3K36-specific methyltransferases, leading to a reduction in all methylation states of H3K36[29]. Similarly, the H3K27M mutation disrupts the repressive H3K27me3 mark, resulting in widespread epigenetic reprogramming and aberrant cellular proliferation or tumorigenesis[30]. In our study, neither H3K36M nor H3K27M mutations were detected in any of the BC tissue samples examined by histological staining. These findings suggest that while H3 mutations play pivotal roles in certain malignancies, their contribution to BC pathogenesis, particularly in the context of our cohort, appears minimal. This highlights the need for further exploration into the unique histone modification patterns that define BC subtypes.

The most substantial finding presented here is that H3K9me2 deposited by G9a serves as an independent predictor, with its histological staining results effectively distinguishing TNBC from other breast cancer subtypes. Furthermore, inhibition of G9a drastically impairs the proliferation of breast cancer cell lines from various subtypes, including luminal A, claudin-low TNBC and basal-like TNBC, underscoring its potential as a therapeutic target across these subtypes. Comparing to H3K9me3 that is often linked to the formation of heterochromatin defining stable and highly repressive chromatin states, H3K9me2 is less stable, associated with dynamic repression[7]. Its writer G9a has broader substrate specificity, targeting both histone and non-histone proteins, such as p53, CDYL, and ERα[31]. G9a is well-recognized as a coactivator in ER-positive breast cancer, where its inhibition reactivates p53 and induces necroptosis, highlighting its dual roles in tumor progression[32]. Mechanistically, G9a directly methylates ERα, thereby enhancing its transcriptional activity on genes that drive cell growth and survival[17]. Experimental depletion of G9a in breast cancer cells and colorectal cancer stem cells has been shown to suppress motility and disrupt ECM organization, underscoring its broader role in cancer cell dynamics[33, 34]. Importantly, to our knowledge, our study is the first transcriptomic analysis profiling TNBC cells after G9a inhibitor treatment, uncovering pathways that overlap but are not identical to RNAseq result from published G9a-knockdown experiments[35]. Our transcriptomic analysis of G9a-inhibited BC cells supports previous findings by revealing G9a’s critical role in maintaining ER activity in ER-positive BC cells and promoting ECM signaling pathways in both ER-positive or - negative BC cells. While ER signaling is specific to ER-positive cells, EMT pathway regulation appears consistent across both ER-positive and ER-negative breast cancer cells, highlighting G9a’s broader influence on cellular processes irrespective of estrogen receptor status[36, 37].

Given G9a’s involvement in estrogen receptor coactivation, it is considered a potential therapeutic target for ER-positive breast cancer. However, its epigenetic role in ER-negative breast cancer remains uncertain, given the absence of ER signaling in these cancers. Notably, the IHC score of H3K9me2 in our cohorts is significantly higher in higher-grade and later-stage tumors compared to healthy tissue, lower-grade, earlier-stage or less aggressive subtypes like luminal A. This observation suggests that H3K9me2 may play a more crucial role in the progression of more aggressive cancers or breast cancer that doesn’t have hormone receptors. Consistent to that, our findings show that G9a inhibition has a more profound inhibitory effect on cancer growth in basal-like TNBC > claudin-low TNBC > luminal A BC, highlighting G9a’s broader influence across different BC subtypes via distinct pathways and mechanisms. Additionally, our results reinforce earlier conclusions that G9a inhibitors induces apoptosis in breast cancer cell lines, with a more pronounced tumor volume reduction in MDA-MB-231 cells compared to MCF-7 cells[37]. Although cell proliferation was impaired, our transcriptomic profiling revealed that G9a inhibition in TNBC cell lines uniquely induces the upregulation of oncogene target genes and the downregulation of genes targeted by tumor suppressors such as SATB1 and TP53[38, 39]. This contrasts with prior studies that reported G9a inhibitors effectively suppress tumor growth in preclinical models, though our findings diverge in terms of the reactivation of silenced tumor suppressor genes[36, 40, 41]. This observation suggests that the heightened sensitivity of ER-negative breast cancer cells to G9a inhibitors may be influenced by the epigenetic changes induced by treatment, rather than reflecting a direct mechanistic basis.

This study, while providing valuable insights, has several limitations that warrant consideration. The relatively small cohort size and lack of follow-up or survival data impede a comprehensive understanding of the prognostic value of histone biomarkers.

Additionally, the scoring methodology, reliant on manual assessment, requires significant labor, experienced technicians, and carries the risk of manual error, emphasizing the need for automation through machine learning-driven imaging technology. Furthermore, the absence of in vivo validation limits the ability to confirm the therapeutic potential of G9a inhibitors, highlighting the need for preclinical studies and further investigation into its dualistic functions in breast cancer subtypes.

## Conclusion

The immunohistochemical profiling and transcriptomic analyses presented in this study identify effective subtype-specific epigenetic targets in various BC subtypes, with a particular focus on TNBC. The findings highlight G9a’s multifaceted role in regulating both hormone-driven pathways and tumor suppressor mechanisms, demonstrating its context-dependent influence. This research sheds light on the epigenetic complexity underlying TNBC and provides valuable insights for developing targeted therapeutic strategies, offering a promising avenue to improve outcomes for patients with this challenging breast cancer subtype.

## Consent for publication

All authors have read the manuscript and are consentaneous for publication.

## Author contributions

XZ conceptualized the study design and the workflow, supervised all aspects of the research, drafted figures and finalized the manuscript. ZH and SZ conducted experiments (except those otherwise noted), processed samples, analyzed data, prepared illustrations, and contributed to manuscript preparation. GS performed IHC staining for the discovery cohort. YC supported the manuscript drafting and provided critical feedback. RC developed the bioinformatics analysis pipeline. MJ, DY, and SZ offered supports on drafting experimental protocols and performing statistical analysis. YX assisted with experiment design, supervised the experiments, and critically reviewed the manuscript.

## Data Availability

Raw and processed files for RNA sequences (FASTQ format) supporting the findings of this study have been deposited in the National Center for Biotechnology Information (NCBI) Gene Expression Omnibus (GEO) under accession number GSE283819.

## Acknowledgments

We extend our gratitude to the commercial organizations and their patient contributors for providing the tissue samples used in this study.

---

**Figure S1.**
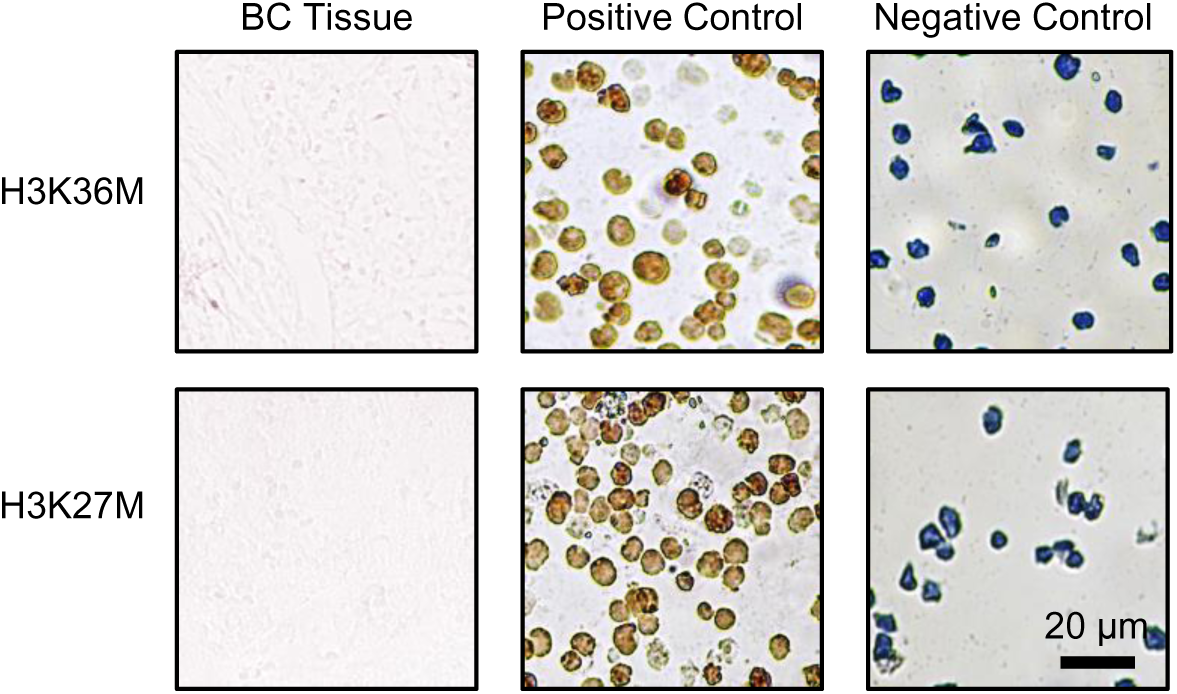

**Figure S2.**
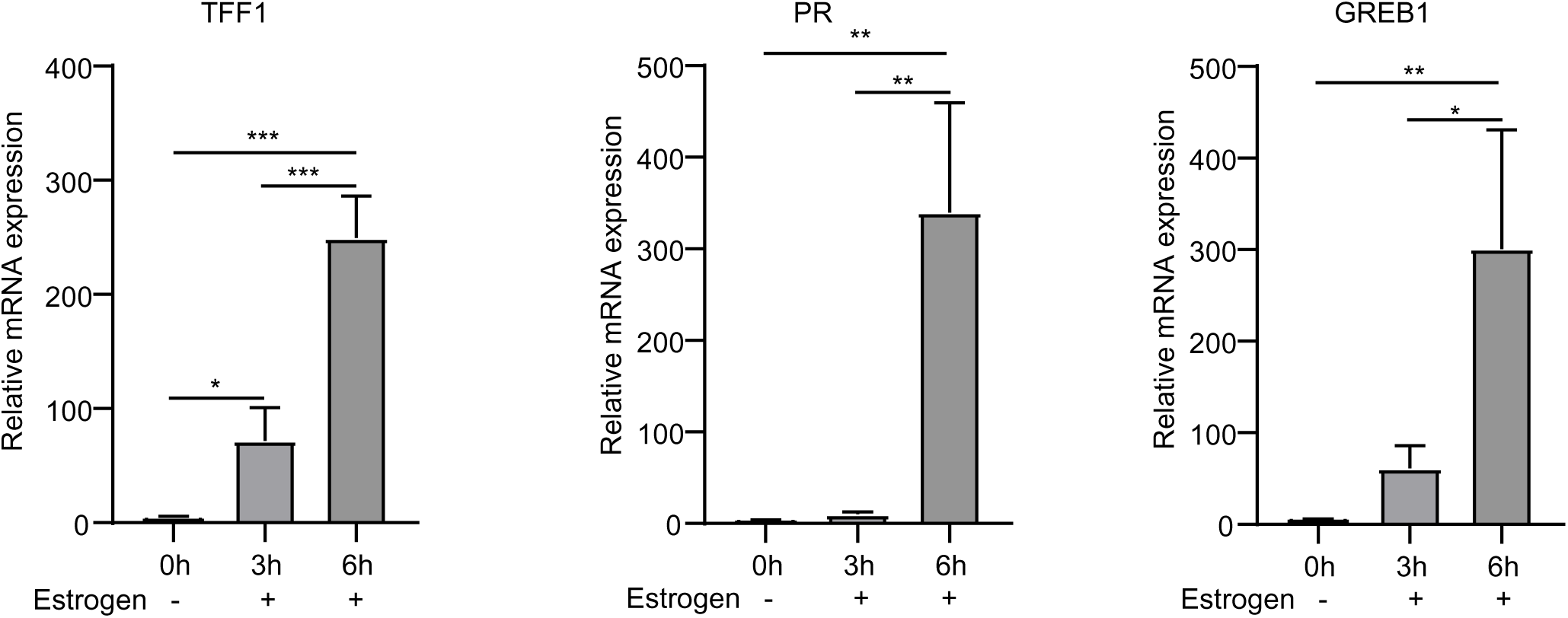

**Figure S3.**
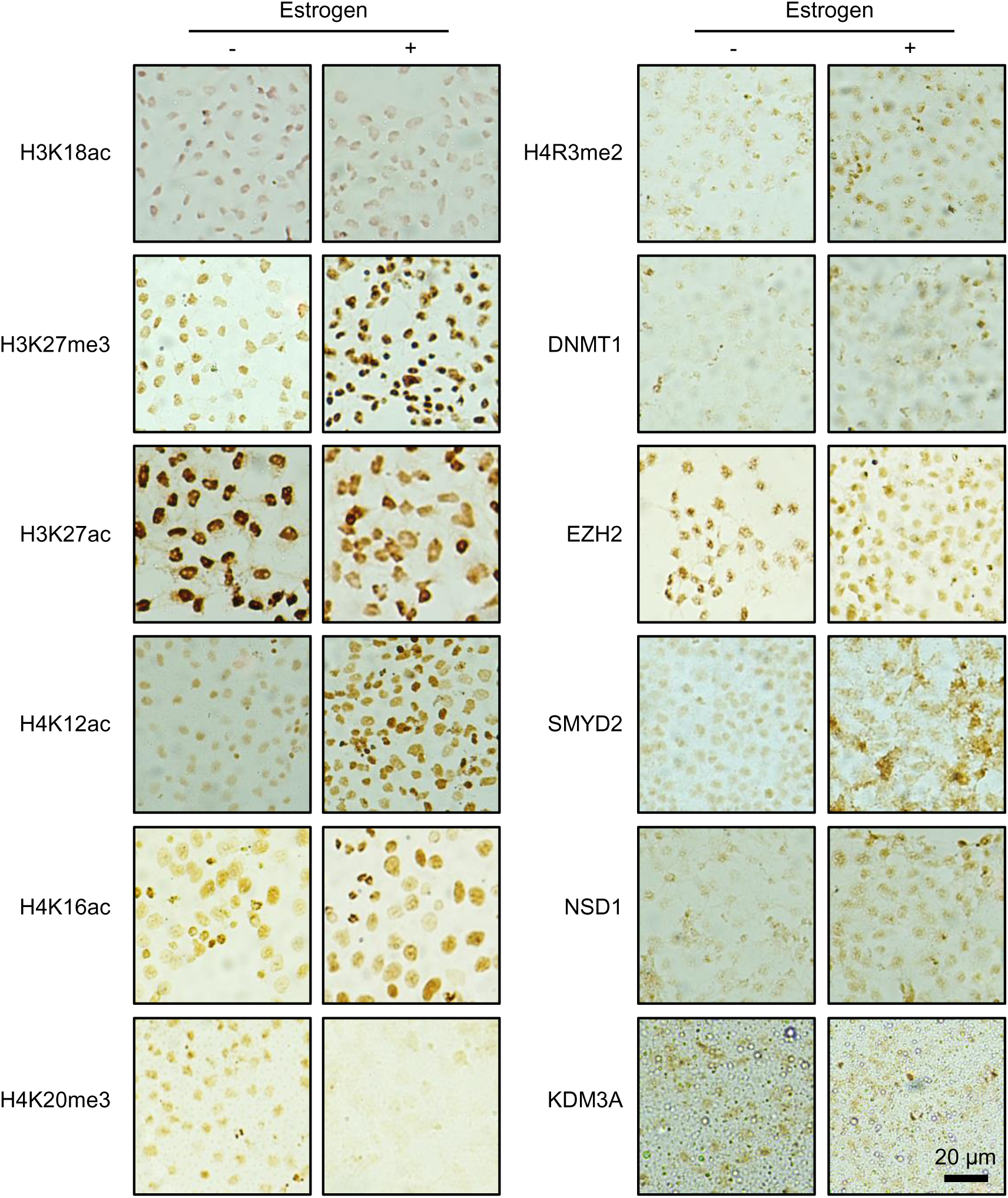

**Figure S4.**
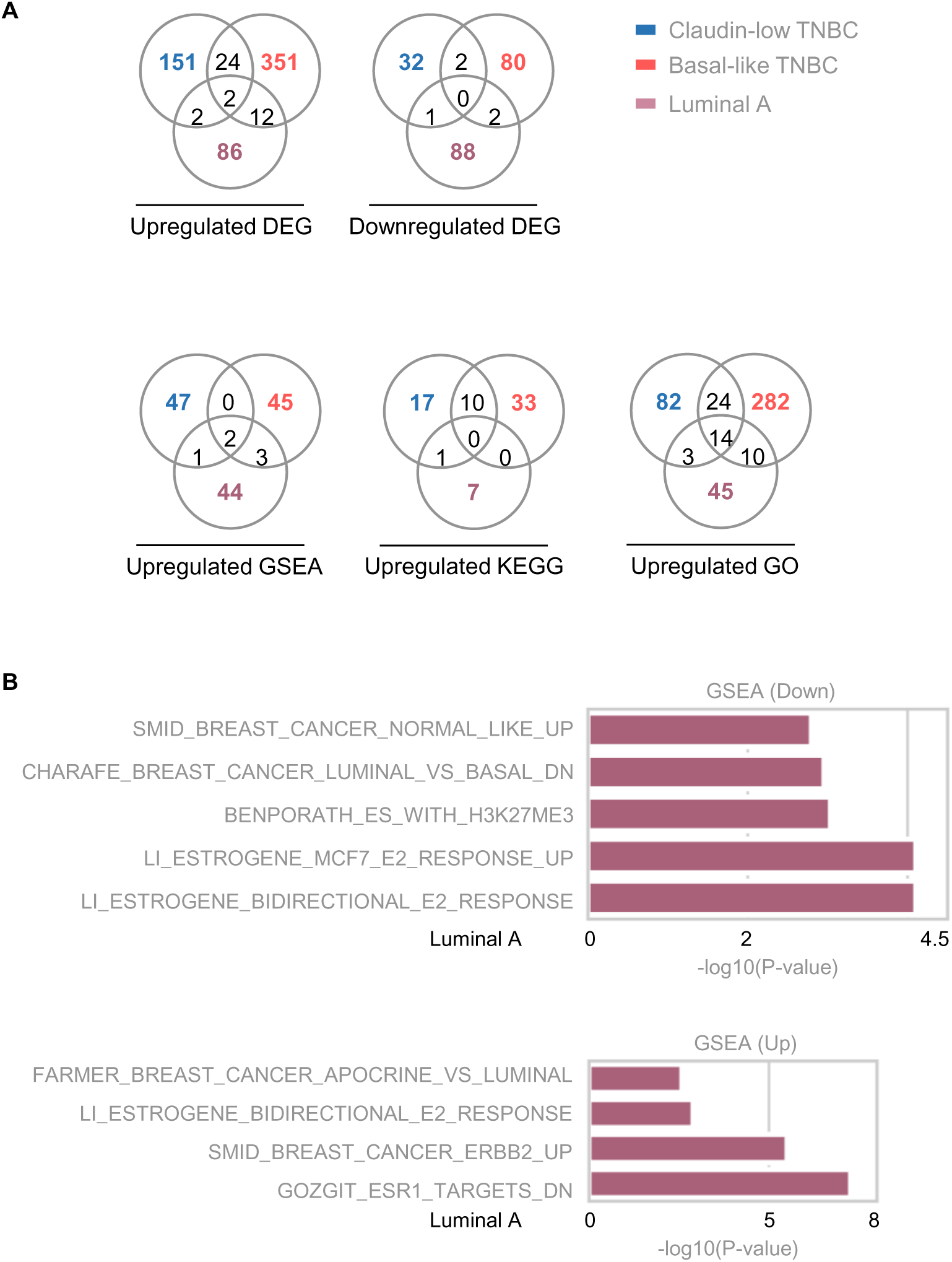

## Reference

1. Bannister, A.J. and T. Kouzarides, Regulation of chromatin by histone modifications. Cell Res, 2011. 21(3): p. 381–95.

2. Yang, Y., M. Zhang, and Y. Wang, The roles of histone modifications in tumorigenesis and associated inhibitors in cancer therapy. J Natl Cancer Cent, 2022. 2(4): p. 277–290.

3. Zhang, X., H. Wen, and X. Shi, Lysine methylation: beyond histones. Acta Biochim Biophys Sin (Shanghai), 2012. 44(1): p. 14–27.

4. Beňačka, R., et al., Classic and New Markers in Diagnostics and Classification of Breast Cancer. Cancers (Basel), 2022. 14(21).

5. Eccles, S.A., et al., Critical research gaps and translational priorities for the successful prevention and treatment of breast cancer. Breast Cancer Res, 2013. 15(5): p. R92.

6. Zhuang, J., et al., Perspectives on the Role of Histone Modification in Breast Cancer Progression and the Advanced Technological Tools to Study Epigenetic Determinants of Metastasis. Front Genet, 2020. 11: p. 603552.

7. Xi, Y., et al., Histone modification profiling in breast cancer cell lines highlights commonalities and differences among subtypes. BMC Genomics, 2018. 19(1): p. 150.

8. Borkiewicz, L., Histone 3 Lysine 27 Trimethylation Signature in Breast Cancer. Int J Mol Sci, 2021. 22(23).

9. Li, W., et al., Targeting Histone Modifications in Breast Cancer: A Precise Weapon on the Way. Front Cell Dev Biol, 2021. 9: p. 736935.

10. Cheng, Y., et al., Targeting epigenetic regulators for cancer therapy: mechanisms and advances in clinical trials. Signal Transduct Target Ther, 2019. 4: p. 62.

11. Marzochi, L.L., et al., Use of histone methyltransferase inhibitors in cancer treatment: A systematic review. Eur J Pharmacol, 2023. 944: p. 175590.

12. White, J., et al., Histone lysine acetyltransferase inhibitors: an emerging class of drugs for cancer therapy. Trends Pharmacol Sci, 2024. 45(3): p. 243–254.

13. Liao, Q., et al., Novel insights into histone lysine methyltransferases in cancer therapy: From epigenetic regulation to selective drugs. J Pharm Anal, 2023. 13(2): p. 127–141.

14. Gold, S. and A. Shilatifard, Epigenetic therapies targeting histone lysine methylation: complex mechanisms and clinical challenges. J Clin Invest, 2024. 134(20).

15. Feng, J. and X. Meng, Histone modification and histone modification-targeted anti-cancer drugs in breast cancer: Fundamentals and beyond. Front Pharmacol, 2022. 13: p. 946811.

16. Zhang, X., et al., Regulation of estrogen receptor α by histone methyltransferase SMYD2-mediated protein methylation. Proc Natl Acad Sci U S A, 2013. 110(43): p. 17284–9.

17. Zhang, X., et al., G9a-mediated methylation of ERα links the PHF20/MOF histone acetyltransferase complex to hormonal gene expression. Nat Commun, 2016. 7: p. 10810.

18. Chen, Y., et al., MicroRNA-133b is regulated by TAp63 while no gene mutation is present in colorectal cancer. Oncol Rep, 2017. 37(3): p. 1646–1652.

19. Fang, Y., et al., Protein expression of ZEB2 in renal cell carcinoma and its prognostic significance in patient survival. PLoS One, 2013. 8(5): p. e62558.

20. Vini, R., et al., 27-Hydroxycholesterol represses G9a expression via oestrogen receptor alpha in breast cancer. J Cell Mol Med, 2023. 27(18): p. 2744–2755.

21. Garcia-Martinez, L., et al., Epigenetic mechanisms in breast cancer therapy and resistance. Nat Commun, 2021. 12(1): p. 1786.

22. Prasanna, T., et al., A Phase 1 Proof of Concept Study Evaluating the Addition of an LSD1 Inhibitor to Nab-Paclitaxel in Advanced or Metastatic Breast Cancer (EPI-PRIMED). Front Oncol, 2022. 12: p. 862427.

23. Noce, B., et al., LSD1 inhibitors for cancer treatment: Focus on multi-target agents and compounds in clinical trials. Front Pharmacol, 2023. 14: p. 1120911.

24. Byun, W.S., et al., Targeting Histone Methyltransferase DOT1L by a Novel Psammaplin A Analog Inhibits Growth and Metastasis of Triple-Negative Breast Cancer. Mol Ther Oncolytics, 2019. 15: p. 140–152.

25. Jeong, G.Y., et al., NSD3-Induced Methylation of H3K36 Activates NOTCH Signaling to Drive Breast Tumor Initiation and Metastatic Progression. Cancer Res, 2021. 81(1): p. 77–90.

26. Grasset, E.M., et al., Triple-negative breast cancer metastasis involves complex epithelial-mesenchymal transition dynamics and requires vimentin. Sci Transl Med, 2022. 14(656): p. eabn7571.

27. Sahu, V. and C. Lu, *Oncohistones:* Hijacking the histone code. Annu Rev Cancer Biol, 2022. 6: p. 293–312.

28. Fang, D., et al., The histone H3.3K36M mutation reprograms the epigenome of chondroblastomas. Science, 2016. 352(6291): p. 1344–8.

29. Rajagopalan, K.N., et al., Depletion of H3K36me2 recapitulates epigenomic and phenotypic changes induced by the H3.3K36M oncohistone mutation. Proc Natl Acad Sci U S A, 2021. 118(9).

30. Harutyunyan, A.S., et al., H3K27M induces defective chromatin spread of PRC2-mediated repressive H3K27me2/me3 and is essential for glioma tumorigenesis. Nat Commun, 2019. 10(1): p. 1262.

31. Zhang, X., Y. Huang, and X. Shi, Emerging roles of lysine methylation on non-histone proteins. Cell Mol Life Sci, 2015. 72(22): p. 4257–72.

32. Mabe, N.W., et al., G9a Promotes Breast Cancer Recurrence through Repression of a Pro-inflammatory Program. Cell Rep, 2020. 33(5): p. 108341.

33. Jin, Y., et al., G9a Knockdown Suppresses Cancer Aggressiveness by Facilitating Smad Protein Phosphorylation through Increasing BMP5 Expression in Luminal A Type Breast Cancer. Int J Mol Sci, 2022. 23(2).

34. Bergin, C.J., et al., G9a controls pluripotent-like identity and tumor-initiating function in human colorectal cancer. Oncogene, 2021. 40(6): p. 1191–1202.

35. Kim, K., et al., RNA-seq based transcriptome analysis of EHMT2 functions in breast cancer. Biochem Biophys Res Commun, 2020. 524(3): p. 672–676.

36. Liu, X.R., et al., UNC0638, a G9a inhibitor, suppresses epithelial-mesenchymal transition-mediated cellular migration and invasion in triple negative breast cancer. Mol Med Rep, 2018. 17(2): p. 2239–2244.

37. Dong, C., et al., G9a interacts with Snail and is critical for Snail-mediated E-cadherin repression in human breast cancer. J Clin Invest, 2012. 122(4): p. 1469–86.

38. Han, H.J., et al., SATB1 reprogrammes gene expression to promote breast tumour growth and metastasis. Nature, 2008. 452(7184): p. 187–93.

39. Silwal-Pandit, L., et al., TP53 mutation spectrum in breast cancer is subtype specific and has distinct prognostic relevance. Clin Cancer Res, 2014. 20(13): p. 3569–80.

40. Zhang, Q., et al., Discovery of novel G9a/GLP covalent inhibitors for the treatment of triple-negative breast cancer. Eur J Med Chem, 2023. 261: p. 115841.

41. Casciello, F., et al., G9a-mediated repression of CDH10 in hypoxia enhances breast tumour cell motility and associates with poor survival outcome. Theranostics, 2020. 10(10): p. 4515–4529.

