## Supplementary material for "Immunohistochemical Profiling of Histone Modification Biomarkers Identifies Subtype-Specific Epigenetic Signatures and Potential Drug Targets in Breast Cancer": legends and tables

**Figure Legends**

**Figure 1. Workflow diagram depicting the identification of histone biomarker signatures in breast cancer.** Histone biomarkers strongly associated with molecular types of breast cancer (BC) were identified through immunohistochemical (IHC) staining in tissue and cell line samples. The functional roles of histone biomarker signatures were further investigated using cell line models treated with small-molecule inhibitor targeting histone methyltransferases.

**Figure 2. Pathological staining and scoring of BC tissue samples from the discovery cohort.** A) The histone biomarkers analyzed in this study are listed. IHC scoring results for the discovery cohort are shown as the percentage of tissue samples with high IHC scores (High-IHC %). B) Low-power IHC images display the expression of four classic BC biomarkers (with nuclear counterstain) and four representative histone biomarkers (without nuclear counterstain). C) High-power IHC images illustrate representative histone biomarker expression at both high and low IHC scores. Staining results are annotated as P (positive for classic biomarkers) and H or L (high or low IHC scores for histone biomarkers).

**Figure 3. Distribution of histone biomarker levels across BC molecular subtypes.** A) Representative H&E and IHC images depicting the four BC molecular subtypes in the discovery cohort, classified based on the results of classic IHC biomarkers, as shown. B) Heatmap and hierarchical clustering of the high-IHC % of each histone biomarkers across the molecular subtypes. Numbers within the colored circles represent the high-IHC % for the corresponding biomarker. Asterisks denote statistical significance of four statistical analyses (Tables 2 &3): 1) distribution of histone biomarkers across 4 molecular subgroups; 2) distribution of histone biomarkers between TNBC vs other molecular subtypes; 3) univariate regression analysis performed for each histone biomarker as a predictor of IHC score groups; 4) multivariate regression for each histone biomarker as an independent predictor. C) Representative IHC images showing the expression of selected histone biomarkers across the molecular subtypes. Staining results are annotated as H (high-IHC score), and L (low-IHC score).

**Figure 4. IHC of TNBC-specific histone biomarker signatures in tissue and cell lines.** A) High-power IHC image representatives of TNBC-specific histone biomarker signatures in adjacent healthy breast tissue (Healthy), luminal A breast cancer tissue and TNBC tissue. B) Immunocytochemical (ICC) staining image representatives of TNBC-specific histone biomarker signatures in luminal A cell lines (MCF-7) without or with estrogen treatment. Staining results are annotated as H or L (high or low-IHC score).

**Figure 5. Correlation Between Histone Biomarker Levels and Tumor Grade/Stage.** (A) Heatmap depicting high-IHC % for selected histone biomarkers across tumor grades and stages. Numbers within the colored circles represent the specific high-IHC % for each biomarker. Only histone biomarkers with significantly different distributions (Table 2) across at least two grade or stage subgroups are shown. (B) Bar plots illustrating normalized high-IHC % of selected histone biomarkers across tumor grades and stages. Normalization was performed using the lowest grade (Grade 2) and stage (N0 or T2) as references. Only histone biomarkers with significantly different distributions across molecular subtypes are displayed.

**Figure 6. Cell proliferation is impaired in TNBC cell lines following G9a inhibition.**

A) Cell viability of three breast cancer cell lines (MCF-7 as luminal A, MDA-MB-231 as claudin-low TNBC, and MDA-MB-468 as basal-like TNBC) under treatment of small-molecule G9a inhibitor UNC0642 was assessed using CCK-8 assay to optimize the treatment conditions. B & C) ICC staining (B) and western blotting (C) were performed to assess the expression of H3K9me2 and other key histone modifications in three cell lines that were treated with UNC0642. D) Cell proliferation of three breast cancer cell lines was assessed using CCK-8 assay.

**Figure 7. Transcriptomic analysis of BC cell lines treated with G9a inhibition**

A) **Volcano plots** showing differentially expressed genes (DEGs) in each cell line after inhibitor treatment, with upregulated and downregulated genes annotated by color. The x-axis represents log2 fold change, and the y-axis indicates -log10 FDR p-value. B) **Venn diagrams** illustrating the overlap of significantly downregulated GSEA term, KEGG and GO pathways among the three cell lines, highlighting shared and cell-line-specific pathways affected by the inhibitor. C-E) **Bar plots** of representative enriched pathways, GO and GSEA terms either down-regulated or up-regulated in BC cell lines as indicated. The x-axis represents the -log10 FDR p-value.

**Supplementary Figure S1.** Two oncohistone mutations were assessed using IHC staining in BC tissues. The LOUCY cell line was used as a positive control for H3K36M and a negative control for H3K27M, while HPB-ALL served as a positive control for H3K27M and a negative control for H3K36M.

**Supplementary Figure S2.** Relative mRNA levels of ER alpha-responsive genes TFF1, PR, and GREB1 were measured by qRT-PCR in MCF-7 cells treated with estrogen for 3 and 6 h. The mRNA level was normalized to GAPDH and retalive levels were calculated using 0 h as reference. Error bars indicate the mean ± s.e.m. of three replicates. Significant fold changes are indicated as follows: *P < 0.05; **P < 0.01; ***P < 0.001 (Student's t-test).

**Supplementary Figure S3.** ICC staining images of additional histone biomarkers in MCF-7 cell lines without or with estrogen treatment.

**Supplementary Figure S4.** A) **Venn diagrams** illustrating the overlap of significantly regulated DEG, GSEA term, KEGG and GO pathways among the three cell lines. B) Bar plots of enriched GSEA terms regulated in luminal A cells (MCF-7), with each bar representing a GSEA term. The x-axis represents the -log10 FDR p-value.

**Table S1** DEG lists identified from breast cancer cell lines treated with G9a inhibition

**Table S1a** List of DEGs identified in MCF-7 cells, n=193

**Table S1b** List of DEGs identified in MDA-MB-231, n=214

**Table S1c** List of DEGs identified in MDA-MB-468, n=473

**Table S2** GSEA identified from breast cancer cell lines treated with G9a inhibition

**Table S2a** Upregulated GSEA identified in MCF-7 cells (p < 0.05), n=50

**Table S2b** Downregulated GSEA identified in MCF-7 cells (p < 0.05), n=50

**Table S2c** Upregulated GSEA identified in MDA-MB-231 cells (p < 0.05), n=50

**Table S2d** Downregulated GSEA identified in MDA-MB-231 cells (p < 0.05), n=43

**Table S2e** Upregulated GSEA identified in MDA-MB-468 cells (p < 0.05), n=50

**Table S2f** Downregulated GSEA identified in MDA-MB-468 cells (p < 0.05), n=50

**Table S3** Pathways identified from breast cancer cell lines treated with G9a inhibition

**Table S3a** Upregulated KEGG pathways identified in MCF-7 cells (p < 0.05), n=8

**Table S3b** Downregulated KEGG pathways identified in MCF-7 cells (p < 0.05), n=11

**Table S3c** Upregulated KEGG pathways identified in MDA-MB-231 cells (p < 0.05), n=28

**Table S3d** Downregulated KEGG pathways identified in MDA-MB-231 cells (p < 0.05), n=10

**Table S3e** Upregulated KEGG pathways identified in MDA-MB-468 cells (p < 0.05), n=43

**Table S3f** Downregulated KEGG pathways identified in MDA-MB-468 cells (p < 0.05), n=4

**Table S3g** Upregulated GO items identified in MCF-7 cells (p < 0.05), n=72

**Table S3h** Downregulated GO items identified in MCF-7 cells (p < 0.05), n=84

**Table S3i** Upregulated GO items identified in MDA-MB-231 cells (p < 0.05), n=123

**Table S3j** Downregulated GO items identified in MDA-MB-231 cells (p < 0.05), n=230

**Table S3k** Upregulated GO items identified in MDA-MB-468 cells (p < 0.05), n=331

**Table S3l** Downregulated GO items identified in MDA-MB-468 cells (p < 0.05), n=87

**Table S4** List of reagents and supplies used in this study

**Table S4a** List of tissue sample codes used in this study

**Table S4b** List of cell lines used in this study

**Table S4c** List of reagents and kits used in this study

**Table S4d** List of antibodies used in this study

**Table S4e** List of PCR primers used in this study

**Table 1. Characteristics of cohort patients and the classic BC biomarker features**

| **Characteristic Features** | | **Discovery Cohort:**  **all BC cases**  **(Specimen N=196)** | **Discovery Cohort: TNBC cases**  **(Specimen N=41)** | **Validation Cohort: TNBC cases**  **(Specimen N=20)** | **P value^*^** |
| --- | --- | --- | --- | --- | --- |
| Mean age (year) | | 50.4 ± 11.0 | 49.2 ± 12.2 | 57.5 ± 12.5 | **0.017** |
| Grade (%) | 1-2 | 0 (0%) | 0 (0%) | 1 (5.0%) | 0.654 |
|  | 2 | 158 (80.6%) | 23 (56.1%) | 9 (45.0%) |  |
|  | 2-3 | 16 (8.2%) | 10 (24.4%) | 6 (30.0%) |  |
|  | 3 | 22 (11.2%) | 8 (19.5%) | 4 (20.0%) |  |
| T stage (%) | T1 | 6 (3.0%) | 0 (0%) | 2 (10.0%) | 0.052 |
|  | T2 | 94 (48.0%) | 20 (48.8%) | 13 (65.0%) |  |
|  | T3 | 58 (29.6%) | 8 (19.5%) | 3 (15.0%) |  |
|  | T4 | 38 (19.4%) | 13 (31.7%) | 2 (10.0%) |  |
| N stage (%) | N0 | 67 (68.4%) | 32 (78.0%) | 17 (85.0%) | 0.546 |
|  | N1 | 46 (23.4%) | 5 (12.2%) | 0 (0%) |  |
|  | N1-2 | 0 (0%) | 0 (0%) | 3 (15.0%) |  |
|  | N2 | 16 (8.2%) | 4 (9.8%) | 0 (0%) |  |
| Tumor stage (%) | I | 4 (2.0%) | 0 (0%) | 2 (10.0%) | 0.083 |
|  | II | 132 (67.3%) | 26 (63.4%) | 15 (75.0%) |  |
|  | III | 60 (30.6%) | 15 (36.6%) | 3 (15.0%) |  |
| ER (%) | Negative | 78 (39.8%) | 41 (100%) | 20 (100%) | NA |
|  | Positive | 118 (60.2%) | 0 (0%) | 0 (0%) |  |
| PR (%) | Negative | 96 (49.0%) | 41 (100%) | 20 (100%) | NA |
|  | Low (<20%) | 18 (9.2%) | 0 (0%) | 0 (0%) |  |
|  | High (≥20%) | 82 (41.8%) |  |  |  |
| HER2 (%) | Negative | 158 (80.6%) | 41 (100%) | 20 (100%) | NA |
|  | Positive | 38 (19.4%) | 0 (0%) | 0 (0%) |  |
| Ki-67 (%) | Low (<14%) | 106 (54.1%) | 17 (41.5%) | 1 (5.0%) | **0.003** |
|  | High (≥14%) | 90 (45.9%) | 24 (58.5%) | 19 (95.0%) |  |

* A t-test was used to compare the mean Age values, while a chi-square test was applied to compare the distribution of TNBC cases across different characteristic groups between the Discovery cohort (N = 41) and the Validation cohort (N = 20).

**Table 2. Comparison of histone biomarker levels across sample subgroups defined by patient characteristics**

| **Histone Biomarkers** | **Statistical significance of the distributions (P value*)** | | | | | | | |
| --- | --- | --- | --- | --- | --- | --- | --- | --- |
|  | **Grade** | **T Stage** | **N Stage** | **Tumor Stage** | **PR** | **Ki-67** | **Molecular Subtypes^#^** | **TNBC^##^** |
| H3K4me2 | **0.009** | 0.068 | **0.001** | 0.073 | 0.619 | **0.000** | **0.014** | 0.374 |
| H3K9me2 | **0.001** | **0.043** | **0.003** | 0.052 | 0.597 | **0.002** | **0.003** | **0.025** |
| H3K9ac | 0.368 | 0.321 | 0.094 | 0.359 | 0.192 | **0.002** | 0.235 | 0.387 |
| H3K18ac | 0.741 | 0.251 | **0.000** | **0.001** | **0.003** | 0.843 | 0.142 | **0.024** |
| H3K27me3 | 0.422 | 0.939 | **0.046** | 0.421 | 0.099 | 0.620 | 0.978 | 0.919 |
| H3K27ac | 0.111 | 0.079 | **0.018** | **0.045** | **0.045** | 0.566 | 0.158 | 0.219 |
| H3K36me2 | 0.077 | 0.431 | 0.094 | 0.406 | 0.473 | **0.006** | **0.022** | **0.010** |
| H3K79me | **0.012** | **0.002** | 0.072 | **0.007** | **0.001** | 0.159 | 0.160 | **0.029** |
| H4K12ac | 0.273 | **0.005** | 0.055 | **0.028** | **0.046** | 0.462 | 0.988 | 0.791 |
| H4K16ac | 0.545 | 0.328 | 0.787 | 0.367 | **0.020** | 0.906 | 0.111 | **0.036** |
| H4K20me3 | **0.007** | 0.225 | **0.032** | **0.031** | **0.015** | 0.101 | 0.058 | 0.055 |
| H4R3me2 | **0.001** | **0.000** | **0.001** | **0.000** | 0.96 | 0.386 | 0.896 | 0.880 |
| DNMT1 | **0.005** | **0.037** | 0.898 | **0.042** | 0.592 | **0.003** | **0.047** | 0.879 |
| G9a | **0.001** | 0.185 | **0.000** | 0.421 | 0.989 | 0.649 | 0.057 | 0.110 |
| EZH2 | **0.012** | 0.073 | **0.008** | **0.019** | 0.267 | **0.012** | 0.062 | 0.502 |
| SMYD2 | 0.508 | **0.000** | 0.758 | **0.001** | 0.627 | **0.010** | 0.152 | 0.294 |
| NSD1 | **0.048** | 0.141 | **0.028** | 0.190 | 0.160 | **0.001** | **0.043** | 0.481 |
| LSD1 | 0.107 | 0.245 | 0.082 | 0.182 | 0.418 | **0.001** | **0.006** | 0.834 |
| KDM3A | 0.466 | **0.000** | 0.395 | **0.000** | 0.165 | 0.287 | 0.109 | 0.904 |

* Chi-square test to compare the distribution of high-IHC % across sample subgroups defined by each characteristic provided in Table 1. Only selected characteristics that show statistically significant differences (P<0.05) of at least one histone biomarker in the grouping of samples are presented here.

^#^ Comparing across four sample subgroups: Lumina A, Lumina B, HER2-enriched and TNBC subtypes.

^##^ Comparing between TNBC and non-TNBC subgroups.

**Table 3. Logistic regression analysis for histone biomarkers associated with individual molecular subtypes**

| **Histone Biomarkers** | **Univariate Regression (P value)** | | | | **Multivariate Regression (P value)** | | | |
| --- | --- | --- | --- | --- | --- | --- | --- | --- |
|  | **LuminalA** | **LuminalB** | **HER2-enriched** | **TNBC** | **LuminalA** | **LuminalB** | **HER2-enriched** | **TNBC** |
| H3K4me2 | **0.003** | **0.016** | 0.833 | 0.375 | **0.002** | 0.160 |  |  |
| H3K9me2 | 0.160 | **0.001** | 0.861 | **0.027** |  | **0.039** |  | **0.012** |
| H3K9ac | 0.224 | 0.058 | 0.762 | 0.389 |  |  |  |  |
| H3K18ac | 0.282 | 0.810 | 0.383 | **0.026** |  |  |  | 0.057 |
| H3K27me3 | 0.933 | 0.781 | 0.701 | 0.919 |  |  |  |  |
| H3K27ac | 0.530 | 0.630 | 0.052 | 0.221 |  |  |  |  |
| H3K36me2 | **0.01** | 0.474 | 0.784 | **0.012** | **0.016** |  |  | **0.013** |
| H3K79me | 0.284 | 0.861 | 0.380 | **0.033** |  |  |  | **0.001** |
| H4K12ac | 0.899 | 0.931 | 0.795 | 0.791 |  |  |  |  |
| H4K16ac | 0.073 | 0.654 | 0.482 | **0.038** |  |  |  | 0.121 |
| H4K20me3 | **0.014** | 0.812 | 0.318 | 0.057 | **0.000** |  |  |  |
| H4R3me2 | 0.951 | 0.590 | 0.519 | 0.880 |  |  |  |  |
| DNMT1 | **0.01** | **0.043** | 0.339 | 0.879 | 0.170 | 0.056 |  |  |
| G9a | 0.519 | 0.281 | 0.367 | **0.013** |  |  |  | 0.191 |
| EZH2 | **0.01** | **0.049** | 0.727 | 0.503 | 0.172 | 0.769 |  |  |
| SMYD2 | 0.170 | **0.048** | 0.502 | 0.295 |  | **0.017** |  |  |
| NSD1 | **0.015** | **0.02** | 0.685 | 0.482 | 0.394 | 0.409 |  |  |
| LSD1 | **0.006** | **0.002** | 0.645 | 0.834 | 0.186 | **0.015** |  |  |
| KDM3A | 0.091 | **0.036** | 0.481 | 0.904 |  | 0.077 |  |  |

**Table 4. TNBC-specific histone biomarker signatures across subtypes and cohorts**

| **High-IHC %** | **H3K9me2** | **H3K36me2** | **H3K79me** | **G9a** |
| --- | --- | --- | --- | --- |
| #1: Discovery Luminal A (N=79) | 37 (46.8%) | 31 (39.2%) | 28 (35.4%) | 9 (11.4%) |
| #2: Discovery TNBC (N=41) | 23 (56.1%) | 28 (68.3%) | 7 (17.1%) | 8 (19.5%) |
| P value (#2 vs. #1)* | **0.010** | **0.003** | **0.036** | 0.226 |
| #3: Validation Cohort (N=20) | 15 (75.0%) | 12 (60.0%) | 6 (30.0%) | 3 (15.0%) |
| P value (#3 vs. #1)* | **0.024** | 0.094 | 0.647 | 0.659 |
| P value (#3 vs. #2)* | 0.153 | 0.522 | 0.247 | 0.667 |

* Chi-square test provided to compare the distribution of high-IHC % across sample subgroups as indicated.
